# Investigating different factors on the outcome of severe Idiopathic Granulomatous Mastitis in 110 women from 2017 to 2021: A cross-sectional study

**DOI:** 10.1101/2024.06.12.24308853

**Authors:** Mahsa Zarpoosh, Zahra Asgari, Parsa Amirian, Tahereh Sadat Mousavi, Arash Golpazir, Mehran Pournazari

## Abstract

**Aim:** Idiopathic Granulomatous Mastitis is a distressing chronic disease without precise etiology and cure. While this disease is rare worldwide, it has been recorded as a prevalent one in Middle Eastern countries; therefore, our aim to investigate the role of different factors on the outcome of the severe form of the disease can shed light on various aspects of the disease.

**Methods:** We retrospectively identified 110 Iranian women, from 2017 to 2021, with severe Idiopathic Granulomatous Mastitis who met the required histological criteria of this benign disease. All patients had developed severe mastitis according to their signs and required treatment. After gathering the information retrospectively, we analyzed them by using statistical tests.

**Results:** In our study, most patients were overweight or obese in their reproductive age with a positive history of pregnancy and breastfeeding. It typically appeared as a painful erythematous mass-like lesion in the upper-outer of the left breast (mean size: 26.75 ± 20.08 mm). Approximately half of the participants experienced extra-mammary manifestations such as arthritis, hair loss, or erythema nodosum. Even though all patients were treated with corticosteroids and Methotrexate or Azathioprine, 32.33% were not completely remitted. Lastly, we realized that there are statistical associations between three variables (family history of Granulomatous Mastitis, the size of the lesions, and the number of recurrences) and the final outcome; all P-values were less than 0.05.

**Conclusion:** The remission rate may decrease when the lesion size exceeds 5 cm, and some hereditary factors may play a role in the response to treatment.

## INTRODUCTION

Idiopathic granulomatous mastitis (IGM) is a rare, non-malignant disease affecting breasts, typically in women, with unknown decisive etiology, first reported by Kessler and Wolloch in 1972 (1). Nonetheless, some factors may play a role in the emergence of the disease, such as autoimmunity conditions, hormonal imbalances, microbiological germs, breast trauma, and over-accumulated milk in the breast (2,3). IGM does not have exclusive presentations, but some non-specific signs, including growing breast mass, edema, erythema, pain, ulcer, fistula, discharge, nipple retraction, and axillary lymphadenitis, are noted in this disease (3).

Furthermore, there are no characteristic signs in ultrasonography, mammography, or magnetic resonance imaging (MRI); it can be diagnosed by excluding other known diseases comprising breast cancer, granulomatous inflammation, or autoimmune diseases (such as sarcoidosis, tuberculosis, and Wegener’s granulomatous), and detecting well-defined non-necrotizing epitheloid granulomas pattern in histological samples (4,5). Granuloma is an aggregation of immune cells in response to cell injury (persistent inflammation, infection, toxin, drugs, or allergic agents) to prevent the spreading; it characteristically is the clump of mononuclear leukocytes (mature macrophages) with or without other inflammatory cell types (6).

To treat or alleviate IGM symptoms, different medications have been used before. Corticosteroids, antibiotics, disease-modifying anti-rheumatic drugs (DMARDs), and surgical procedures (abscess drainage or breast-conserving surgery) are helpful treatments that can be administered solely or combined (7).

Although IGM is a rare condition all over the world, it seems to be more prevalent in Asian countries, especially in women with predisposing genetic and environmental factors (8,9). Numerous cases were reported in Iran, which may show the probable relation between the Iranian race and the occurrence of IGM (3,7,9-14); However, none of these studies were performed in the western part of the country. To investigate the probable connections between different factors and final outcomes, we have evaluated 110 women with severe IGM in detail, which has not been performed before.

## MATERIALS AND METHODS

### Study population

Our study is a retrospective cross□sectional study of 110 female patients of any age with severe IGM in the western part of Iran from 2017 to 2021 with a two-year post-diagnosis period. We retrospectively collected data via the medical files. After a primary physical examination and requesting ultrasonography or laboratory tests by a rheumatologist, an appropriate type of biopsy was done for each patient to exclude the breast cancer and confirm the IGM disease. In addition, the severity criteria were based on clinical/ultrasonography signs (presence of fistula, abscess, or collection bigger than 5cm) and requisite treatments (DMARDs with/without resection or abscess drainage).

Due to the incomplete subsidence in some cases, we deemed it necessary to evaluate the effect of different factors on the outcome of this disease; therefore, we divided patients into two groups by the outcome (remission and under-treatment groups).

## Data collection

We designed a dedicated checklist with the following information: age, Body Mass Index (BMI), gravidity, parity, abortion, pregnancy/lactating/menopausal state, the time interval between the recent childbirth/abortion and the end of lactation to diagnosis, duration of lactation, history/duration of oral contraceptive (OCP) consumption, family history of IGM/ breast cancer, Rheumatic and non-Rheumatic comorbidities, drug history, IGM signs, ultrasonography findings, biopsy type, laterality and location, number and size of the lesion, triggers of onset and exacerbation, other organs involvement, medical treatments and surgical procedures, final outcome, number of recurrences, recurrence stimulants, positive laboratory tests, and Bone mineral density (BMD) tests; then, we created database in Microsoft Excel (Microsoft Corp.).

### Statistical analysis

The data was analyzed through STATA-17 (StataCorp. 2021. Stata Statistical Software: Release 17: StataCorp LLC.). The results were collected as means with standard deviation and percentages. We used the Chi-square independence test to prove the associations between variables and the final outcome in each group; then, we drew charts and graphs. (P-value < 0.05 was considered statistically significant.)

### Ethics approval

This study was conducted according to the principles of the Declaration of Helsinki. It was approved by the Ethics Committee of Kermanshah University of Medical Sciences (IR.KUMS.MED.REC.1401.182).

## RESULTS

The mean age of the patients was 34.98 ± 7.21 years with a range of 22 to 65 years, and the mean BMI was 28.98 ± 3.90 with a range of 22.65 to 40.62 kg/m2 (Table 1). While the mean gravidity and parity were 1.94 ± 0.89 and 1.89 ± 0.90, respectively, 3 cases (2.72%) did not experience pregnancy (Table 1). IGM symptoms started during pregnancy in 5 patients (4.54%), seven during lactation (6.36%), and two in the menopausal period (1.81%) (Table 1). Additionally, six patients (5.45%) had a positive history of abortion. The mean years of the interval between the recent childbirth/or abortion to IGM diagnosis was 5.31 ± 5.22, with a range of 0 to 38 years (Table 1). Although the mean duration of lactation in our cases was 37.26 ± 23.07 months, six women (5.45%) did not report any lactation history. Also, the mean years of interval between the end of lactation to IGM diagnosis was 4.17 ± 5.42 (maximum 36 years) (Table 1). Additionally, 16 patients (14.54%) had a positive family history of breast cancer, and 21 patients (19.9%) had a positive family history of IGM, which can be found in detail in Table 1. The mean duration of OCP consumption was 17.71 ± 36.85 months, ranging from 0 to 192 months (Table 1). Twenty-two patients (20%) had at least one non-rheumatic comorbidity, such as thyroid dysfunction, type 2 diabetes mellitus, hypertension, and polycystic ovary syndrome (Table 1), and 27 participants (24.54%) reported rheumatic/musculoskeletal diseases including arthritis, arthralgia, discopathy, low back pain, legs pain, and Sarcoidosis; which are mentioned in Table 1, in detail. Sixteen cases (14.54%) had a positive history of anti-rheumatic drug consumption, such as NSAIDs (non-steroidal anti-inflammatory drugs), corticosteroids, or DMARDs (disease-modifying anti-rheumatic drugs) (Table 1).

**TABLE 1.** Different characteristics of the IGM and their correlation to the outcome (N = 110)

| Variables | Total patients<br>(n=110) | Mean ± SD<br>(range) | Final outcome | | P-value |
| --- | --- | --- | --- | --- | --- |
|  |  |  | Remission<br>(n=74) | Under treatment<br>(n=36) |  |
| Age (years) | | 34.98 ± 7.21<br>(22 – 65) | | | 0.439 |
| $\leq 20$ | 0 | | 0 | 0 | |
| 21-30 | 36 |  | 24 | 12 |  |
| 31-40 | 51 |  | 37 | 14 |  |
| 41-50 | 17 |  | 10 | 7 |  |
| 51-60 | 1 |  | 0 | 1 |  |
| $> 60$ | 1 | | 1 | 0 | |
| BMI (kg/m<sup>2</sup>) | | 28.98 ± 3.90<br>(22.65 – 40.62) | | | 0.389 |
| $< 25$ | 9 | | 4 | 5 | |
| 25-30 | 29 |  | 20 | 9 |  |
| $> 30$ | 19 | | 11 | 8 | |
| Gravidity (n) | | 1.94 ± 0.89<br>(0 – 4) | | | 0.991 |
| Positive | 106 |  | 71 | 35 |  |
| Negative | 3 |  | 2 | 1 |  |
| Parity (n) | | 1.89 ± 0.90<br>(0 – 4) | | | 0.728 |
| Positive | 105 |  | 70 | 35 |  |
| Negative | 4 |  | 3 | 1 |  |
| History of abortion (n) | | 0.05 ± 0.22<br>(0 – 1) | | | 0.071 |
| Positive | 6 |  | 2 | 4 |  |
| Negative | 103 |  | 71 | 32 |  |
| Status |  |  |  |  |  |
| Pregnant | 5 |  | 4 | 1 | 0.535 |
| Lactescent | 8 |  | 5 | 3 | 0.765 |
| Menopause | 2 |  | 1 | 1 | 0.599 |
| None | 95 |  | 64 | 31 |  |
| The time interval between the recent childbirth/abortion to diagnosis (years) | | 5.31 ± 5.22<br>(0 – 38) | | | 0.706 |
| $< 5$ | 59 | | 38 | 21 | |
| 5-10 | 36 |  | 26 | 10 |  |
| $> 10$ | 8 | | 5 | 3 | |
| History of lactation (months) | | 37.26 ± 23.07<br>(0 – 96) | | | 0.535 |
| Positive | 99 |  | 66 | 33 |  |
| Negative | 5 |  | 4 | 1 |  |
| The time interval between the end of lactation to diagnosis (years) | | 4.17 ± 5.42<br>(0 – 36) | | | 0.322 |
| $< 5$ | 71 | | 46 | 25 | |
| 5-10 | 15 |  | 12 | 3 |  |
| $> 10$ | 8 | | 4 | 4 | |

TABLE 1 (continued)
| Variables | Total patients<br>(n=110) | Mean ± SD | Final outcome | | P-value |
| --- | --- | --- | --- | --- | --- |
|  |  |  | Remission<br>(n=74) | Under treatment<br>(n=36) |  |
| Family history of breast cancer |  |  |  |  | 0.360 |
| Negative | 94 |  | 64 | 30 |  |
| First-degree relative | 2 |  | 0 | 2 |  |
| Second-degree relative | 6 |  | 4 | 2 |  |
| Third-degree relative | 4 |  | 3 | 1 |  |
| Fourth degree relative | 4 |  | 3 | 1 |  |
| Family history of IGM |  |  |  |  | **0.038** |
| Negative | 89 |  | 61 | 28 |  |
| First-degree relative | 12 |  | 10 | 2 |  |
| Second-degree relative | 8 |  | 2 | 6 |  |
| Third-degree relative | 1 |  | 1 | 0 |  |
| History of OCP Consumption<br>(months) | | 17.71 ± 36.85<br>(0 – 192) | | | 0.275 |
| Positive | 44 |  | 32 | 12 |  |
| Negative | 53 |  | 33 | 20 |  |
| Non-RMD comorbidity |  |  |  |  | 0.841 |
| Negative | 88 |  | 57 | 31 |  |
| Hypothyroidism | 9 |  | 6 | 3 |  |
| Hyperthyroidism | 4 |  | 3 | 1 |  |
| Hyperprolactinemia | 3 |  | 3 | 0 |  |
| DM type2 | 6 |  | 4 | 2 |  |
| HTN | 2 |  | 1 | 1 |  |
| CVD | 4 |  | 3 | 1 |  |
| PCOs | 5 |  | 3 | 2 |  |
| Anemia | 2 |  | 1 | 1 |  |
| History of cancer | 3 |  | 3 | 0 |  |
| Lichen planus | 1 |  | 1 | 0 |  |
| Vitiligo | 1 |  | 0 | 1 |  |
| RMD comorbidity |  |  |  |  | 0.184 |
| Negative | 83 |  | 60 | 23 |  |
| Arthritis/Arthralgia | 17 |  | 8 | 9 |  |
| Discopathy/Low back pain | 10 |  | 7 | 3 |  |
| Legs pain | 3 |  | 1 | 2 |  |
| Sarcoidosis | 1 |  | 1 | 0 |  |
| History of anti-rheumatic drugs |  |  |  |  | 0.344 |
| Negative | 94 |  | 66 | 28 |  |
| NSAIDs | 7 |  | 4 | 3 |  |
| Corticosteroids | 8 |  | 4 | 4 |  |
| DMARDs | 3 |  | 1 | 2 |  |
| Laterality |  |  |  |  | 0.312 |
| Left | 60 |  | 39 | 21 |  |
| Right | 34 |  | 26 | 8 |  |
| Bilateral | 16 |  | 9 | 7 |  |
| Number of involved sites |  |  |  |  | 0.455 |
| Single | 48 |  | 33 | 15 |  |
| Multiple | 25 |  | 15 | 10 |  |

TABLE 1 (continued)
| Variables | Total patients<br>(n=110) | Mean ± SD | Final outcome | | P-value |
| --- | --- | --- | --- | --- | --- |
|  |  |  | Remission<br>(n=74) | Under treatment<br>(n=36) |  |
| Location |  |  |  |  | 0.583 |
| Inferior | 12 |  | 6 | 6 |  |
| Inferolateral | 9 |  | 6 | 3 |  |
| Inferomedial | 9 |  | 7 | 2 |  |
| Lateral | 15 |  | 10 | 5 |  |
| Medial | 12 |  | 9 | 3 |  |
| Superior | 7 |  | 4 | 3 |  |
| Superolateral | 22 |  | 17 | 5 |  |
| Superomedial | 4 |  | 4 | 0 |  |
| Size of mass-like lesions (mm) | | 26.75 ± 20.08<br>(6 – 140) | | | 0.028 |
| <50 | 55 |  | 40 | 15 |  |
| >50 | 18 |  | 8 | 10 |  |
| Triggers of onset (self-report) |  |  |  |  | 0.582 |
| None | 18 |  | 11 | 7 |  |
| Covid-19 vaccination | 16 |  | 9 | 7 |  |
| Covid-19 infection | 2 |  | 2 | 0 |  |
| Stress | 5 |  | 4 | 1 |  |
| Breast trauma | 4 |  | 3 | 1 |  |
| Metal contact | 1 |  | 1 | 0 |  |
| Milk accumulation | 1 |  | 0 | 1 |  |
| Triggers of exacerbation (self-report) |  |  |  |  | 0.744 |
| None | 12 |  | 7 | 5 |  |
| Covid-19 vaccination | 3 |  | 2 | 1 |  |
| Covid-19 infection | 4 |  | 4 | 0 |  |
| Stress | 55 |  | 37 | 18 |  |
| Breast trauma | 1 |  | 0 | 1 |  |
| Dehydration | 5 |  | 4 | 1 |  |
| Milk accumulation | 2 |  | 1 | 1 |  |
| Excitant foods | 4 |  | 2 | 2 |  |
| Menstruation | 5 |  | 4 | 1 |  |
| Cold foods | 9 |  | 6 | 3 |  |
| Other organs involvement |  |  |  |  | 0.844 |
| None | 49 |  | 31 | 18 |  |
| Arthritis/Arthralgia | 16 |  | 13 | 3 |  |
| Bone pain | 13 |  | 8 | 5 |  |
| Hair loss | 14 |  | 9 | 5 |  |
| Erythema nodosum | 5 |  | 3 | 2 |  |
| Paresthesia | 3 |  | 2 | 1 |  |
| positive lab tests |  |  |  |  | 0.198 |
| CRP & ESR | 24 |  | 10 | 14 |  |
| Vitamin D deficiency | 12 |  | 8 | 4 |  |
| Prolactin | 4 |  | 3 | 1 |  |
| ACE | 2 |  | 2 | 0 |  |

TABLE 1 (continued)
| Variables | Total patients<br>(n=110) | Mean ± SD | Final outcome | | P-value |
| --- | --- | --- | --- | --- | --- |
|  |  |  | Remission<br>(n=74) | Under treatment<br>(n=36) |  |
| BMD |  |  |  |  | 0.717 |
| Normal | 20 |  | 10 | 10 |  |
| Osteopenia | 3 |  | 1 | 2 |  |
| Osteoporosis | 3 |  | 2 | 1 |  |
| Final treatment |  |  |  |  | 0.740 |
| Corticosteroids | 110 |  | 74 | 36 |  |
| DMARDs |  |  |  |  |  |
| Methotrexate | 105 |  | 70 | 35 |  |
| Azathioprine | 5 |  | 4 | 1 |  |
| Anti-TNF | 3 |  | 1 | 2 |  |
| Colchicine | 110 |  | 74 | 36 |  |
| Cabergoline | 56 |  | 34 | 22 |  |
| Antibiotics therapy |  |  |  |  | 0.873 |
| Yes | 13 |  | 9 | 4 |  |
| No | 97 |  | 65 | 32 |  |
| Surgery |  |  |  |  | 0.269 |
| None | 86 |  | 57 | 29 |  |
| Breast-conserving surgery | 13 |  | 11 | 2 |  |
| Open Drainage | 11 |  | 6 | 5 |  |
| Recurrences (n) | | 0.30 ± 0.53<br>(0 – 3) | | | **0.043** |
| Non-recurrent | 88 |  | 55 | 33 |  |
| One time | 20 |  | 18 | 2 |  |
| Two times | 1 |  | 1 | 0 |  |
| Three times | 1 |  | 0 | 1 |  |
| Recurrence stimulants (self-report) |  |  |  |  | 0.571 |
| Non-compliance |  |  |  |  |  |
| Feeling better | 10 |  | 9 | 1 |  |
| Side effects | 4 |  | 2 | 2 |  |
| COVID-19 infection | 3 |  | 2 | 1 |  |
| COVID-19 vaccination | 3 |  | 2 | 1 |  |
| Major stress | 2 |  | 2 | 0 |  |
| Local breast trauma | 2 |  | 2 | 0 |  |
| Lactation | 1 |  | 1 | 0 |  |

Abbreviations: BMI, body mass index; OCP, oral contraceptive pills; NSAIDs, non-steroidal anti-inflammatory drugs; DMARDs, disease-modifying anti-rheumatic drugs; RMD, rheumatic and musculoskeletal diseases; DM, diabetes mellitus; HTN, hypertension; CVD, cardiovascular disease; PCOs, polycystic ovary syndrome; CRP, C-reactive protein; ESR, erythrocyte sedimentation rate; ACE, angiotensin-converting enzyme; BMD, bone mineral density; anti-TNF, anti-tumor necrosis factor.
All significant P-values are bold.

The basis of diagnosis in all patients was the detection of granuloma patterns in histological samples obtained via excisional biopsy, core needle biopsy, fine needle aspiration, or combined procedures (Figure 1-A). In clinical examination, mass/masses (86.36%), pain (81.82%), erythema (70.91%), nipple discharge (50.91%), edema (45.45%), fever (29.09%), fistula (16.36%) and pruritus (9.09%) were the common signs of IGM, which are noted in Figure 1-B. Furthermore, most of the patients underwent ultrasonography, and prevalent findings were documented as collection/collections (46.36%), mastitis (26.36%), axillary lymphadenitis (26.36%), mass/masses (17.27%), heterogenous fibro glandular tissue (20%), fistula (13.67%) and abscess (9.09%) (Figure 1-C). As mentioned, most of the patients experienced mass-like lesions; these findings were named masses or collections in ultrasonographic findings, with a mean size of 26.75 ± 20.08 mm (73 lesions), ranging from 6 to 140 mm (Table 1). While the left breast was more affected (54.55%) than the right breast (30.90%), 16 women experienced IGM symptoms on both sides (14.55%) (Table 1). IGM signs were present in different sites of the breast; for example, 24.44% of the lesions or other signs located in superolateral (upper-outer) part, and the rest located in lateral (16.67%), medial (13.33%), inferior (13.33%), inferolateral (10%), inferomedial (10%), Superior (7.78%), and superomedial (4.44%) parts (Table 1). Moreover, 34.25% of cases (25 out of 73) suffered from multiple lesions (Table 1).

**FIGURE 1.**
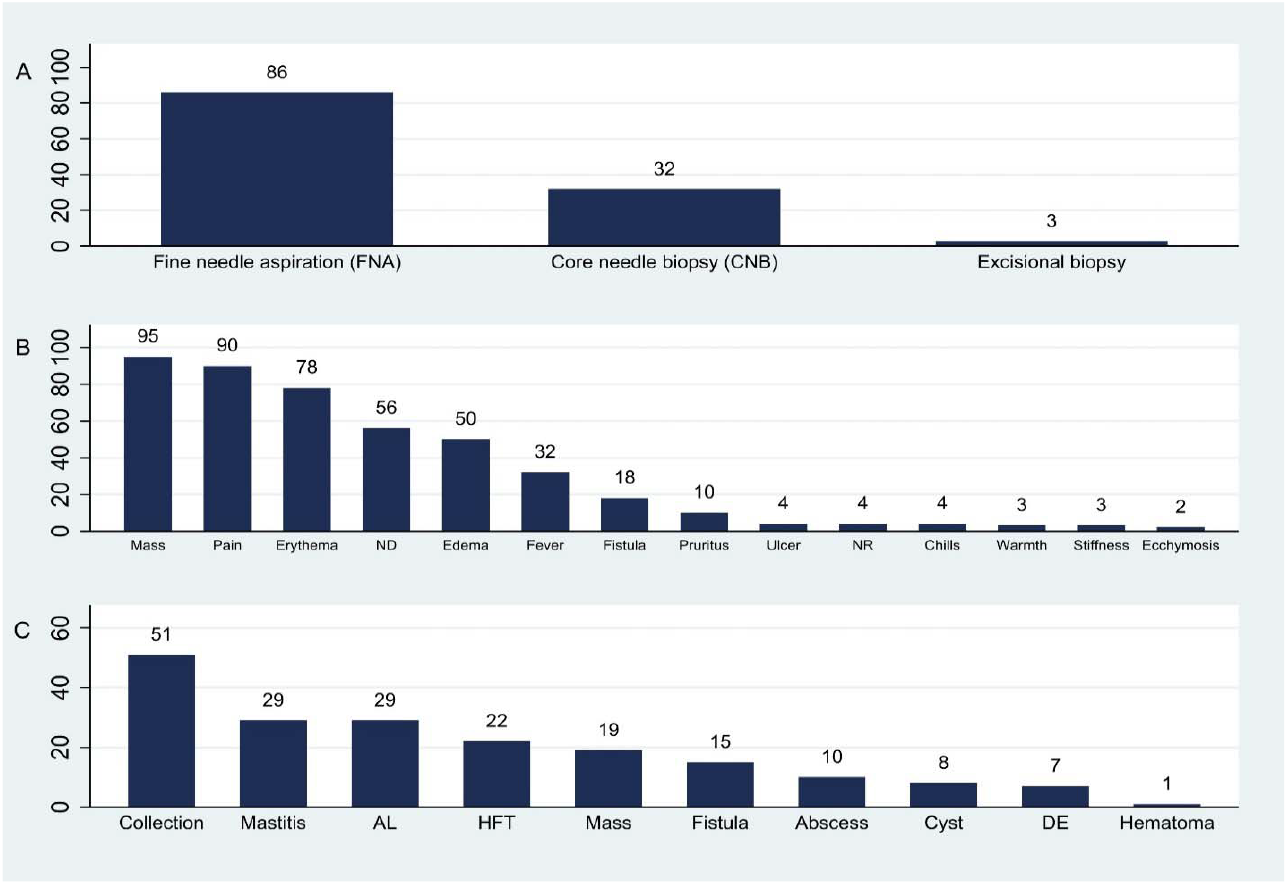
Different biopsy types (A), clinical manifestations of granulomatous mastitis (B), and findings in ultrasonography (C). Abbreviations: ND nipple discharge; NR nipple retraction; AL axillary lymphadenitis; HFT heterogeneous fibro glandular tissue; DE ductal ectasia.

Some patients (26.36%) declared a stimulant factor as the trigger for IGM onset, and 80% of them declaimed a booster factor as the trigger for IGM exacerbation; among causes of onset, Covid-19 vaccination with 14.54% (of all cases) and among causes of exacerbation, stress with 50% (of all cases), were the most frequent ones; other factors are mentioned in Table 1. Moreover, concurrent symptoms, consisting of Arthritis/Arthralgia in 14.54% of patients, bone pain in 11.81%, hair loss in 12.72%, erythema nodosum in 4.54%, and paresthesia in 2.72% were observed (Table 1).

Para-clinical examinations such as laboratory and bone mineral density tests were performed, and 21.81% of cases had high ESR (erythrocyte sedimentation rate) (> 10 mm/hour) with elevated CRP (C - reactive protein) (> 3 mg/dl), and 5.45% suffered from osteopenia or osteoporosis; detailed information can be found in Table 1.

Due to the severity of IGM in our cases, all patients underwent potent treatments, including corticosteroids, DMARDs (Methotrexate in non-pregnant women and Azathioprine in pregnant women), and Colchicine. In addition, 11.81% had breast-conserving surgery, and 10% experienced open abscess drainage (Table 1). Antibiotic therapy was administered in 11.81% of the cases, and anti-TNF (tumor necrosis factor) therapy was prescribed in 2.72% (Table 1). Additionally, Cabergoline was prescribed for patients with nipple discharge (50.91%) (Table 1).

All patients were observed for two years after IGM diagnosis and starting the treatment; at the end of the 24th month, remission was achieved in 67.27% (74 out of 110) of cases successfully, and 32.33% (36 out of 110) were treated partially so needed to continue the treatment (Table 1). Besides, 20% of all patients (22 out of 110) experienced at least one recurrence during this period, and in 86.36% of cases remission was finally achieved (19 out of 22), but not achieved in 13.64% (3 out of 22); one patient had three recurrences, and another one had 2, so the total number of recurrences became 25. Reasons for relapse, based on self-report, were as follows: non-compliance (56%), COVID-19 infection (12%), COVID-19 vaccination (12%), stress (8%), local trauma (8%), and lactation (4%) (Table 1).

Chi-squared test for different variables was performed; after the analysis, it was revealed that there are statistical correlations between the family history of IGM and the outcome of IGM (P-value = 0.038), the size of the mass-like lesions and the outcome (P-value = 0.028), and the number of recurrences and the outcome (P-value = 0.043) (Table1).

## DISCUSSION

Idiopathic granulomatous mastitis (IGM) is an infrequent condition, with an estimated incidence of 2.4 per 100,000 women throughout the world, which is more frequent in Middle Eastern countries such as Iran and Turkey (8,15). Previous studies showed IGM with a wide range of severity (3,5). In this study, we analyzed the data of 110 patients with the severe form of the disease in the western part of Iran; to the best of our knowledge, it has not been done before.

We have concluded that IGM commonly affects overweight or obese women in their reproductive age with a positive history of pregnancy and breastfeeding. Most of the patients suffered from a painful and erythematous mass-like lesion (with a size smaller than 5 cm). The left breast’s upper outer (superolateral) was the usual site of the lesion. Even though most participants did not mention comorbidity, about half of them stated simultaneous extra-mammary involvement. The above findings were also previously shown in other studies (3,11,14-21).

A large number of our cases experienced an exacerbating factor during treatment (stress was the most remarkable one), which has not been investigated before. In a study by Rakhshan et al. and a survey by Kaviani et al., the family history of breast cancer in IGM patients was 38.3% and 44.8%, respectively (3,11); however, in our study, the family history of breast cancer was 14.54%, we also investigated the family history of IGM which was 19.9%. Unlike other studies, we only included patients with the severe form of the disease, which led to some different reports in treatments and outcomes; for instance, the recurrence rate of IGM was 11.2% and 12.6% in Deng et al. and Yılmaz et al. studies (18,19), but this rate was a bit higher in our study population (20%); additionally, in a systematic review of 71 studies, by Fattahi et al, the recurrence rate was 17.8% in 4735 patients (21). Moreover, corticosteroids with DMARD were administered in all cases, which was not the case in similar studies. Although our patients had severe IGM, only 21.81% underwent aggressive treatments such as resection or drainage.

In this study, we explored the feasible role of various factors on final outcomes in the severe form of IGM. We realized that there are statistical correlations between three factors (family history of IGM, size of the mass-like lesions, and the number of recurrences) and the final outcome (P-value < 0.05) (Table 1). Nevertheless, there were no statistical relations between the following factors and the final outcome: age, BMI, gravidity, parity, abortion, pregnancy/lactating/menopausal state, the time interval between the recent childbirth/abortion, the time interval between the end of lactation to diagnosis, duration of lactation, history/duration of oral OCP consumption, family history of breast cancer, Rheumatic and non-Rheumatic comorbidities, drug history, laterality and location, number of the lesions, triggers of onset and exacerbation, other organs involvement, treatments, recurrence stimulants, positive laboratory tests, and BMD test (P-value > 0.05) (Table 1).

During two years after diagnosis, all patients were treated with corticosteroids, DMARDs, and colchicine; at the end of this period, 67.27% (74 out of 110) were treated successfully (remission group), but 32.33% (36 out of 110) still needed to continue the treatment (under-treatment group). In the first group, 82.43% did not report any family history of IGM, but 17.57% had a positive family history (first-degree: 13.51%, second-degree: 2.70%, third-degree: 0.90%); on the other hand, in the second group, 77.78% did not state any family history of IGM, and 22.22% had a positive family history (first-degree: 5.55%, second-degree: 16.67%). Due to our findings, patients with a positive family history of IGM respond harder to the treatment; therefore, we assume that some hereditary factors may play a role in the pathogenicity of IGM (Figure 2-A)(21,22). While the mean size of mass-like lesions was 26.75 ± 20.08 in all patients, 40% of lesions in the under-treatment group and 16.67% in the remission group, were greater than 5cm, which suggests that giant lesions may respond to the treatment, with more difficulty (Figure 2-C).

**FIGURE 2.**
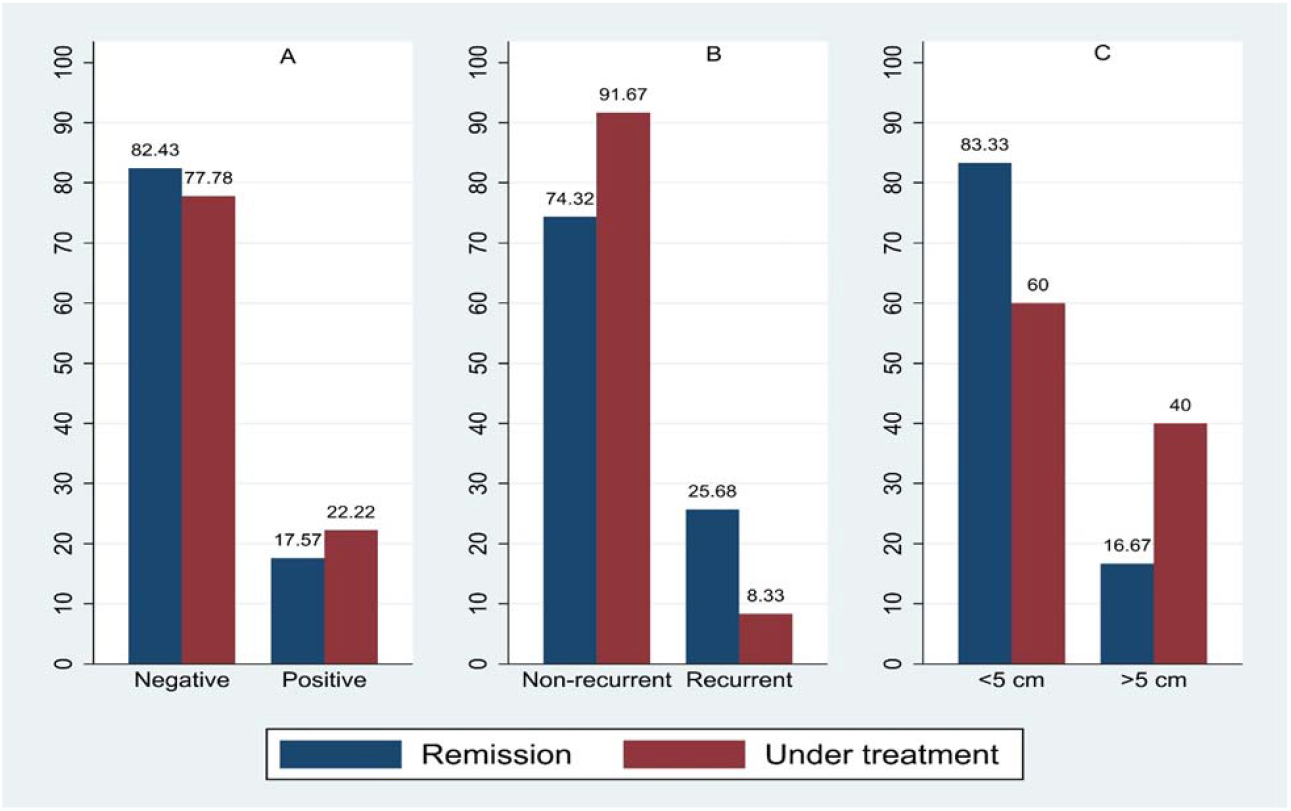
Comparison of family history of IGM (A), recurrence of IGM (B), and the size of mass-like lesions (C) between two categories of remission and under-treatment, based on percentage.

Unexpectedly, 25.68% of patients who had complete remission had at least one recurrence in the two-year post-diagnosis period, but this rate was lower in the under-treatment group (8.33%) (Figure 2-B). We suppose that two reasons may play a role; firstly, most patients in the remission group confirmed non-adherence to the treatment as they felt better in the post-diagnosis period, whereas women in the under-treatment group had higher rates of compliance to the medications (Table 1); secondly, two years is a limited time for evaluating the recurrence, it may change in more extended periods.

Besides all the new data and investigations, our study had some limitations; some data were not found in the medical documents, and patients were unavailable due to the COVID-19 pandemic. A larger study population with a long-term follow-up is suggested.

## CONCLUSION

We concluded that there may be an inverse relationship between remission rate and mass-like lesion size, and some hereditary factors may also cause difficulties in response to the treatment. The recurrence rate was higher in patients with complete remission, which may be due to inadequate follow-up time or non-adherence to treatment after feeling better.

## Data Availability

The data supporting the present study's findings are available from the corresponding author upon reasonable request.

## Author Contributions

Mahsa Zarpoosh: data analysis, manuscript writing.

Zahra Asgari: data collection, proposal writing.

Parsa Amirian: data analysis, visualization.

Tahereh Sadat Mousavi: data collection.

Arash Golpazir: conceptualization, validation.

Mehran Pournazari: supervision, review & editing.

## REFERENCES

1. Kessler, E., & Wolloch, Y. (1972). Granulomatous mastitis: a lesion clinically simulating carcinoma. American journal of clinical pathology, 58(6), 642–646. 10.1093/ajcp/58.6.642

2. Altintoprak, F., Kivilcim, T., & Ozkan, O. V. (2014). Aetiology of idiopathic granulomatous mastitis. World Journal of Clinical Cases: WJCC, 2(12), 852. 10.12998/wjcc.v2.i12.852

3. Kaviani, A., Vasigh, M., Omranipour, R., Mahmoudzadeh, H., Elahi, A., Farivar, L., & Zand, S. (2019). Idiopathic granulomatous mastitis: Looking for the most effective therapy with the least side effects according to the severity of the disease in 374 patients in Iran. The breast journal, 25(4), 672–677. 10.1111/tbj.13300

4. Manogna, P., Dev, B., Joseph, L. D., & Ramakrishnan, R. (2020). Idiopathic granulomatous mastitis—our experience. Egyptian Journal of Radiology and Nuclear Medicine, 51(1), 1–8. 10.1186/s43055-019-0126-4

5. Yin, Y., Liu, X., Meng, Q., Han, X., Zhang, H., & Lv, Y. (2022). Idiopathic granulomatous mastitis: etiology, clinical manifestation, diagnosis and treatment. Journal of Investigative Surgery, 35(3), 709–720. 10.1080/08941939.2021.1894516

6. Shah, K. K., Pritt, B. S., & Alexander, M. P. (2017). Histopathologic review of granulomatous inflammation. Journal of clinical tuberculosis and other Mycobacterial Diseases, 7, 1–12. 10.1016/j.jctube.2017.02.001

7. Shojaee, L., Rahmani, N., Moradi, S., Motamedi, A., & Godazandeh, G. (2021). Idiopathic granulomatous mastitis: challenges of treatment in iranian women. BMC surgery, 21, 1–7. 10.1186/s12893-021-01210-6

8. Wolfrum, A., Kümmel, S., Theuerkauf, I., Pelz, E., & Reinisch, M. (2018). Granulomatous mastitis: a therapeutic and diagnostic challenge. Breast care, 13(6), 413–418.. 10.1159/000495146

9. Alikhassi, A., Azizi, F., & Ensani, F. (2020). Imaging features of granulomatous mastitis in 36 patients with new sonographic signs. Journal of Ultrasound, 23, 61–68. 10.1007/s40477-019-00392-3

10. Sari, F., Raei, N., Najafi, S., Haghighat, S., Moghadam, S., & Olfatbakhsh, A. (2022). Idiopathic Granulomatous Mastitis: An institutional Experience from a Referral Center: IGM-diagnostics and treatment approach. Archives of Breast Cancer, 9(3-SI), 301–308. 10.32768/abc.202293SI301-308

11. Rakhshan, A., Akbari, A., Ahadi, M., Zham, H., Moradi, A., Toudeshki, K. K., & Bashiri, S. (2023). Clinicopathological Evaluation of Idiopathic Granulomatous Mastitis: A Retrospective Analysis of Sixty Women at Shohada-e-Tajrish Hospital from 2010 to 2019. International Journal of Cancer Management, (In Press). 10.5812/ijcm-139543

12. Omranipour, R., Mohammadi, S. F., & Samimi, P. (2013). Idiopathic granulomatous lobular mastitis-report of 43 cases from Iran; introducing a preliminary clinical practice guideline. Breast care, 8(6), 439–443. 10.1159/000357320

13. Aghajanzadeh, M., Hassanzadeh, R., Sefat, S. A., Alavi, A., Hemmati, H., Delshad, M. S. E., … & Massahniya, S. (2015). Granulomatous mastitis: presentations, diagnosis, treatment and outcome in 206 patients from the north of Iran. The Breast, 24(4), 456–460. 10.1016/j.breast.2015.04.003

14. Azizi, A., Prasath, V., Canner, J., Gharib, M., Sadat Fattahi, A., Naser Forghani, M., … & Habibi, M. (2020). Idiopathic granulomatous mastitis: Management and predictors of recurrence in 474 patients. The breast journal, 26(7), 1358–1362. 10.1111/tbj.13822

15. Martinez□Ramos, D., Simon□Monterde, L., Suelves□Piqueres, C., Queralt□Martin, R., Granel□Villach, L., Laguna□Sastre, J. M., … & Escrig□Sos, J. (2019). Idiopathic granulomatous mastitis: A systematic review of 3060 patients. The breast journal, 25(6), 1245–1250. 10.1111/tbj.13446

16. Benson, J. R., & Dumitru, D. (2016). Idiopathic granulomatous mastitis: presentation, investigation and management. Future Oncology, 12(11), 1381–1394. 10.2217/fon-2015-0038

17. Yuan, Q. Q., Xiao, S. Y., Farouk, O., Du, Y. T., Sheybani, F., Tan, Q. T., … & Wu, G. S. (2022). Management of granulomatous lobular mastitis: an international multidisciplinary consensus (2021 edition). Military medical research, 9(1), 1–15. 10.1186/s40779-022-00380-5

18. Deng, Y., Xiong, Y., Ning, P., Wang, X., Han, X. R., Tu, G. F., & He, P. Y. (2022). A case management model for patients with granulomatous mastitis: a prospective study. BMC Women’s Health, 22(1), 1–12. 10.1186/s12905-022-01726-w

19. Yılmaz, T. U., Gürel, B., Güler, S. A., Baran, M. A., Erşan, B., Duman, S., & Utkan, Z. (2018). Scoring idiopathic granulomatous mastitis: an effective system for predicting recurrence?. European journal of breast health, 14(2), 112. 10.5152/ejbh.2018.3709

20. Fattahi, A. S., Amini, G., Sajedi, F., & Mehrad-Majd, H. (2023). Factors Affecting Recurrence of Idiopathic Granulomatous Mastitis: A Systematic Review. The Breast Journal, 2023. 10.1155/2023/9947797

21. Sheybani, F., Naderi, H., Gharib, M., Sarvghad, M., & Mirfeizi, Z. (2016). Idiopathic granulomatous mastitis: Long-discussed but yet-to-be-known. Autoimmunity, 49(4), 236–239. 10.3109/08916934.2016.1138221

22. Zhu, Q., Wang, L., & Wang, P. (2021). The identification of gene expression profiles associated with granulomatous mastitis. Breast Care, 16(4), 319–327. 10.1159/000507474

